# Layer-Resolved Spatial Transcriptomic Profiling Reveals Distinct Fibrotic and Muscularizing Submucosal Niches in Fibrostenotic Crohn’s Disease

**DOI:** 10.64898/2026.09.14.26363080

**Authors:** Xianyong Gui, Guangxu Jin, David L. Caudell

## Abstract

**Background and Aims:** Fibrostenotic stricture is a severe complication of Crohn’s disease (CD) that affect up to 50% of patients over their lifetime, causing irreversible narrowing of the intestinal lumen and bowel obstruction, but the molecular programs that distinguish stenotic from non-stenotic inflammation and the relative contributions of fibrosis and smooth-muscle remodeling remain incompletely characterized. We aimed to dissect bowel-wall compartment-specific and histology-defined remodeling programs in fibrostenotic CD.

**Methods:** Matched full-thickness terminal ileal tissues from 4 patients with fibrostenotic CD were sampled from stenotic stricture (SS), non-stenotic inflamed bowel (NS), and non-inflamed bowel (NI). Histomorphology-guided NanoString GeoMx whole-transcriptome profiling was performed across mucosa, muscularis mucosae, submucosa, and muscularis propria. GeoMx data were processed by standard software provided by GeoMx platform. DESeq2 models included patient identity as a blocking factor, and the analysis was performed in 74 histologically valid regions of interest (ROIs) representing full spectrum of CD histopathologies. DEGs were defined as adjusted P < .05 and |log2FC| >= 1.0.

**Results:** Out of 18,677 genes analyzed in each of the ROIs, direct SS-versus-NS comparison identified 0 significant DEGs in mucosa, 18 in muscularis mucosae, 43 in submucosa, and 0 in muscularis propria. NS-versus-NI submucosa showed 132 DEGs (131 increased, 1 decreased), dominated by humoral/plasma-cell genes with a modest stromal-remodeling component. SS-versus-NI submucosa showed 144 DEGs (138 increased, 6 decreased) with coordinated contractile, extracellular-matrix/remodeling and cell-matrix adhesion programs. Muscularized versus fibrotic SS submucosa showed 219 DEGs, with marked enrichment of contractile and remodeling genes in muscularized regions. Excluding 5 follicle-containing submucosal ROIs markedly altered the NS comparison but not the core SS fibromuscular program. Exploratory follicular-niche analyses showed that lymphoid follicle-rich (LFR) submucosa was lymphoid/B-cell enriched relative to lymphoid follicle-null (LFN) areas; SS-LFR nevertheless acquired a stronger fibroblast/ECM program relative to NS-LFR, while CCL19 was the only significant LFR-versus-LFN DEG within SS.

**Conclusions:** Fibrostenotic CD is characterized by spatially concentrated submucosal fibromuscular remodeled inflammatory program. Non-stenotic submucosa is heterogeneous and may already show regional stromal remodeling, whereas established stenosis displays a broader and more robust ECM-contractile-adhesion phenotype with molecularly distinct fibrotic and muscularized niches.

## Introduction

Fibrostenotic stricture is a major and clinically significant complication of Crohn’s disease (CD), that frequently results in irreversible bowel obstruction requiring endoscopic intervention and/or surgical resection. Although anti-inflammatory and biologic therapies can control active inflammation, established structural strictures remain difficult to reverse, and no approved pharmacologic therapy is available.^1-3^

The traditional model of Crohn’s stricture formation has emphasized extracellular-matrix (ECM) deposition and fibrosis, which has led ongoing pharmaceutic studies targeting intestinal fibrosis. Histopathologic studies, however, have shown that the submucosal muscularization and smooth-muscle hyperplasia/hypertrophy are prominent structural components of fibrostenosing bowel.^4-7^ These observations support a broader mechanism of fibrostenosis including transmural mesenchymal and fibromuscular remodeling rather than collagen accumulation alone.

Recent single-cell and spatial transcriptomic studies have begun to resolve the cellular heterogeneity of CD strictures. Single-cell RNA sequencing performed on full-thickness biopsy samples identified fibroblast heterogeneity and suggested that many stricture-selective changes were localized to the mucosa/submucosa rather than the muscularis propria.^8^ Combined single-cell and spatial studies subsequently identified IgG-positive plasma cells, PECAM, inflammatory fibroblasts, collagen-high fibroblasts, and spatially organized multicellular networks in strictures.^9-12^ Other studies identified GREM1 and related stromal genes as stricture-associated biomarkers candidates,^13^ while high-resolution spatial profiling studies identified myofibroblast-dominated fibrotic niches and submucosal muscularization.^14-18^ Despite these advances, several questions remain unresolved. First, it is difficult to separate molecular changes associated with chronic inflammation from those specifically associated with stenosis. Second, bowel-wall layers may contribute unequally to fibrostenosis. Third, histologically fibrotic and muscularized submucosal regions may represent different molecular states rather than different degrees of one process. Finally, spatial heterogeneity within non-stenotic inflamed submucosa may obscure early or regionally restricted remodeling programs.

We therefore performed histomorphology-guided stage- and layer-resolved GeoMx whole-transcriptome profiling of matched full-thickness terminal ileal tissues of segmental stenotic (SS), adjacent inflamed but non-stenotic (NS), and noninflamed (NI) regions from the same patients. Extensive regions of interest (ROIs) were selected to represent a wide spectrum of Crohn’s histopathology. We hypothesized that fibrostenotic stricture would be primarily associated with a spatially concentrated submucosal fibromuscular program and that fibrosis and muscularization would constitute related but molecularly distinguishable remodeling states.

## Materials and Methods

### Study Cohort and Tissue Selection

The cohort included four patients with established fibrostenotic CD who underwent terminal ileal resection during 2023-2025 at Wake Forest Baptist Medical Center. They were retrospectively selected from pathology database based on the adequacy of original diagnostic tissue sampling, including 2 women and 2 men; mean age was 39 years, and mean disease duration was 13.25 years; all patients had received biologic therapy before surgery (Remicade, Humira, Humira and Entyvio, or Cimzia). Twelve representative full-thickness formalin-fixed, paraffin-embedded (FFPE) tissue samples of terminal ileum, 3 for each patient, were selected by an experienced gastrointestinal pathologist (XG) from the archived diagnostic surgical pathology materials.

### Composite Blocks Construction and Spatial Profiling

For each patient, three different regions from the same resection specimen were selected based on confirmatory histopathology review of diagnostic slides: stenotic stricture (SS), defined as the epicenter and maximally thickened portion of the segmental stenotic stricture identified in the surgical specimen; non-stenotic inflamed bowel (NS), defined as adjacent inflamed but non-strictured/non-thickened bowel; and non-inflamed bowel (NI), defined as histologically unremarkable region from the same resected bowel and showed no apparent microscopic inflammation or other abnormalities, generally at or next to the surgical margin. The most representative full-thickness areas were dissected from the original SS, NS, and NI FFPE blocks and re-embedded into a single patient-specific composite block that allowed the three different bowel regions of the same patient be studied simultaneously to facilitate matched spatial profiling. Figure 1 illustrates the composite block construction.

**Figure 1.**
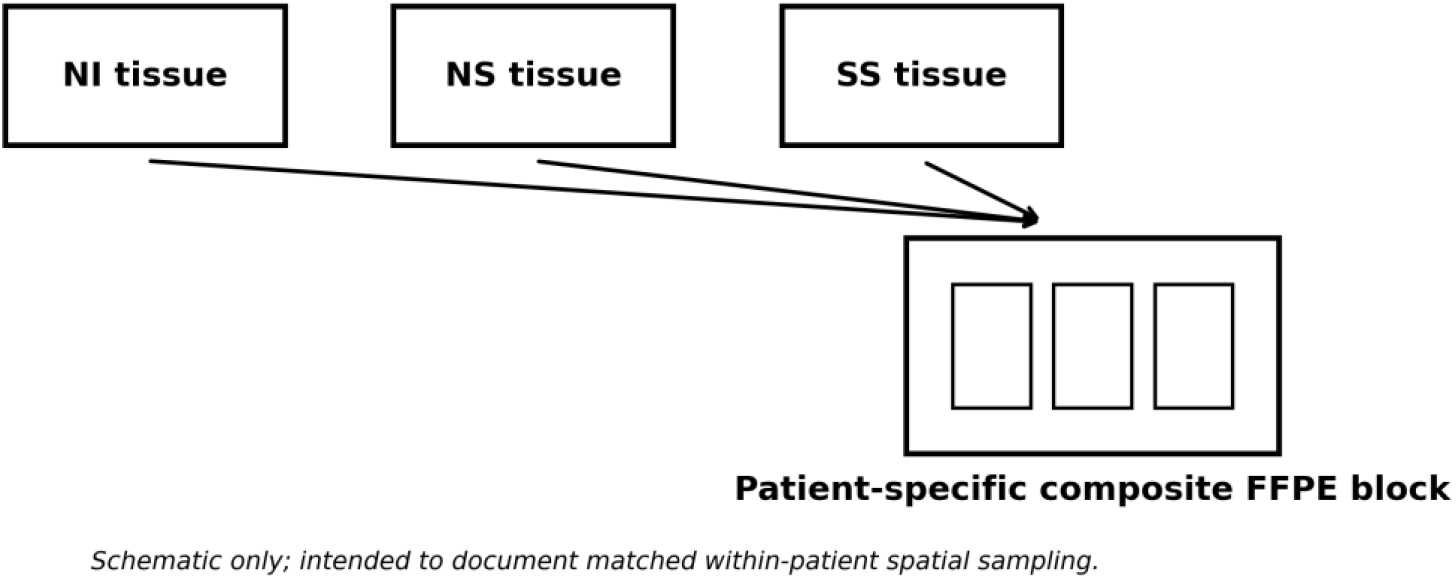
Matched three full-thickness terminal ileal tissue sections representing NI, NS, and SS regions were lined side by side to construct a composite tissue block. Patient-specific composite FFPE blocks were then profiled using GeoMx DSP/WTA with morphology-guided ROI selection across bowel-wall layers.

Spatial transcriptomic studies were performed using the NanoString GeoMx Digital Spatial Profiler (DSP) with the Human Whole Transcriptome Atlas (WTA).

Morphologic characterization incorporated anti-alpha-smooth muscle actin (α-SMA, Novus Biologicals, NBP2-33006AF532) to identify smooth-muscle cells and myofibroblastic cells, anti-fibroblast activation protein (FAP, Abcam, ab311827) to identify activated fibroblastic/myofibroblastic populations, anti-Calponin (Abcam, abab197640) as an additional smooth-muscle/activated myofibroblastic marker, and SYTO13 for nuclei. Immunofluorescence patterns were interpreted together with corresponding hematoxylin-eosin (H&E) histomorphology. Plasma cells and lymphocytes were readily recognizable on H&E cytomorphology alone.

### Region-of-Interest (ROIs) Selection

ROIs were selected using conventional histomorphology and morphology-marker expression to capture distinct histopathologic compartments and remodeling phenotypes, as shown in Table 1. At least six ROI categories were evaluated: (1) inflamed mucosa; (2) inflamed submucosa without apparent fibromuscular remodeling; (3) markedly fibrotic submucosa(defined by predominantly dense fibrosis composed of collagen-rich stroma and fibroblast proliferation, while scant smooth muscle cells were present); (4) submucosa demonstrating prominent smooth-muscle hyperplasia/muscularization (defined by significant areas of submucosal space essentially replaced by cohesive smooth muscle cells while a mild collagen deposition may be mixed); (5) muscularis mucosae demonstrating thickening and loose/dissection with prominent downward extension; and (6) thickened muscularis propria demonstrating smooth-muscle hyperplasia and/or hypertrophy. Associated inflammation was permitted within the latter remodeling compartments. Corresponding compartments in normal area of the bowel from the same patients were used as controls. 74 ROIs in total were analyzed, including at least 18 ROIs from each patient, 18 from each compartment, and 18 from each group.

**Table 1.**
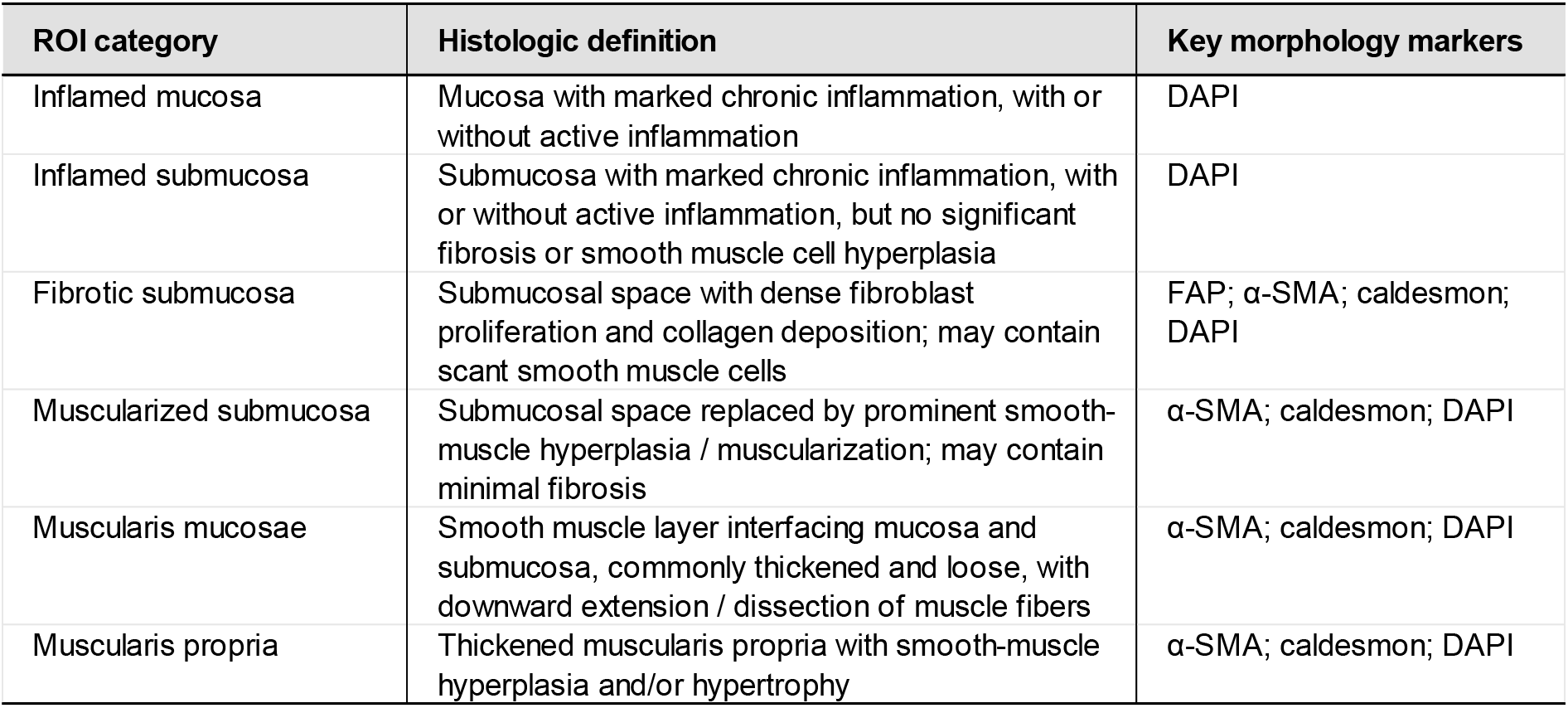
ROIs Definitions.

| ROI category | Histologic definition | Key morphology markers |
| --- | --- | --- |
| Inflamed mucosa | Mucosa with marked chronic inflammation, with or without active inflammation | DAPI |
| Inflamed submucosa | Submucosa with marked chronic inflammation, with or without active inflammation, but no significant fibrosis or smooth muscle cell hyperplasia | DAPI |
| Fibrotic submucosa | Submucosal space with dense fibroblast proliferation and collagen deposition; may contain scant smooth muscle cells | FAP; α-SMA; caldesmon; DAPI |
| Muscularized submucosa | Submucosal space replaced by prominent smooth-muscle hyperplasia / muscularization; may contain minimal fibrosis | α-SMA; caldesmon; DAPI |
| Muscularis mucosae | Smooth muscle layer interfacing mucosa and submucosa, commonly thickened and loose, with downward extension / dissection of muscle fibers | α-SMA; caldesmon; DAPI |
| Muscularis propria | Thickened muscularis propria with smooth-muscle hyperplasia and/or hypertrophy | α-SMA; caldesmon; DAPI |

Oligonucleotide tags corresponding to the WTA probes were photocleaved from each selected spatial region, collected, sequenced, and mapped back to the corresponding ROI.

For the submucosal analysis, all histologically valid ROIs were included initially. Among 59 submucosal ROIs, 5 contained prominent lymphoid follicles: 3 NS ROIs from one patient and 2 SS ROIs from a different patient. Because these follicle-predominant ROIs were patient-clustered rather than independently distributed across the cohort, follicle status was not treated as an independent biological factor in the primary analysis but was further analyzed separately.

### Bioinformatic and Statistical Analysis

GeoMx DSP raw count files (DCC), probe kit configuration files, and sample annotations were imported into R using the GeomxTools Bioconductor package as a NanoStringGeoMxSet object. Sample-, slide-, ROI-, and AOI/segment-level metadata were retained through preprocessing. Zero counts were shifted by one, and quality control was performed at segment and probe levels. AOIs with low sequencing saturation and probes flagged as outliers were removed. QC-passing probe counts were aggregated to gene-level target counts. Third-quartile (Q3) normalization was used for visualization and exploratory analyses.

Differential expression analysis was performed on the QC-filtered gene-level count matrix using DESeq2 negative-binomial generalized linear models. Patient identity (reassigned with a research case number) was included as a blocking factor and pathologist-defined tissue/disease group was the variable of interest. DESeq2 median-of-ratios size factors were estimated internally. Wald-test P values were adjusted for multiple comparisons using the Benjamini-Hochberg procedure. Genes with a Benjamini-Hochberg-adjusted P value < .05 and an absolute log2 fold change (|log2FC|) >= 1.0 were considered significantly differentially expressed.

Pathway enrichment of significant genes was assessed against Reactome, KEGG, Gene Ontology Biological Process, and MSigDB Hallmark gene-set libraries using Enrichr. CONVERGE-AI GeoMx was used as an interactive reproducibility/accessibility layer over the statistical pipeline to facilitate independent re-execution and review of differential-expression and pathway analyses.

### LLM-guided DEG analysis for pathologists (CONVERGE-AI GeoMx)

The experimental design decisions made by expert pathologists defining which of the hundreds of ROIs and AOI segments enter each comparison are the critical determinant of downstream analytical validity. In practice, such design-driven differential expression analyses must be repeated hundreds of times across comparison configurations, and in the traditional workflow each iteration requires a hand-off from pathologist-defined study design to a bioinformatics team for re-execution, making comprehensive GeoMx analysis prohibitively time-consuming. To eliminate this bottleneck, we leveraged an LLM-guided analysis platform, CONVERGE-AI GeoMx, which enables pathologists to specify, execute, and review differential expression and pathway analyses directly through a natural-language interface layered over the statistical pipeline described above (GeomxTools preprocessing, Q3 normalization, DESeq2 negative-binomial models with Benjamini-Hochberg correction, and Enrichr pathway enrichment). Our pathologists applied CONVERGE-AI GeoMx to hundreds of GeoMx DEG analysis designs, shortening the overall analysis cycle from months under the traditional design-then-hand-off workflow to days, while preserving analytical rigor through fixed, version-controlled statistical methods and ensuring transparency and reproducibility, as every analysis remains independently re-executable and reviewable. CONVERGEAI GeoMx is an enterprise product developed in The JIN-AI Laboratory and commercialized by JINAI L.L.C. (https://www.jinlabai.net/).

### Sensitivity Analysis of Lymphoid-Follicle-Predominant Submucosal ROIs

To assess the robustness of the submucosal findings to regional ROI composition, differential-expression analyses were repeated after excluding the 5 follicle-predominant ROIs considering the predominantly dense aggregates of lymphoid cells in composition as a possible confounding factor. Because all 3 NS follicle-predominant ROIs happened to be taken from one patient and formed almost the entire ROIs in submucosa in that sample, while both SS follicle-predominant ROIs from another patient, this analysis was interpreted as a sensitivity analysis to ROI composition and patient representation rather than as a direct test of lymphoid-follicle biology.

### Exploratory Spatial Analysis of Lymphoid-Follicle-Rich and Lymphoid-Follicle-Null Submucosa

To further characterize regional submucosal heterogeneity, exploratory differential-expression analyses compared lymphoid-follicle-rich (LFR) with lymphoid-follicle-null (LFN) submucosal ROIs, examined SS versus NS within LFR submucosa, and compared LFR with LFN within SS. The same DESeq2 framework was used. For descriptive reporting of significant DEGs in these exploratory analyses, adjusted P < .05 and |log2FC| >= 1.0 were applied. Because the number of LFR ROIs was limited and follicular ROIs were not evenly distributed across patients, these analyses were interpreted as hypothesis-generating spatial comparisons rather than evidence of a causal effect of lymphoid follicles.

### Ethics

This study was approved by the Wake Forest University Health Sciences Institutional Review Board (IRB No. 00074626 - Amendment 64), with waiver-of-consent status granted.

## Results

### Common Mucosal Immune Response Shared by Both Stenotic and Non-Stenotic Bowel

Both SS and NS mucosa across all four patient samples showed extensive transcriptional alterations relative to NI. SS versus NI contained 252 significant DEGs (56 increased, 196 decreased), whereas NS versus NI contained 425 (50 increased, 375 decreased). As shown in Figure 2 and Table 2, prominent increases included immunoglobulin genes, particularly IGHG1-4, indicating plasma-cell-associated humoral immune activation; while prominent decreases involved epithelial/metabolic and mucosal-homeostasis genes including FABP6, GSTA1, UGT2A3, and KCNJ13. Direct SS-versus-NS analysis identified no significant mucosal DEGs, indicating that the dominant mucosal transcriptional response reflects shared CD inflammatory and epithelial biology rather than a stenosis-specific program.

**Table 2.** Key mucosal genes in SS versus NI.

| Gene | log2FC (SS vs NI) | padj | Direction | Biologic Significance |
| --- | --- | --- | --- | --- |
| IGHG3 | +6.22 | 7.86E-16 | Up | Strong immunoglobulin heavy chain signal → local B-cell / plasma cell activity |
| IGHG2 | +6.12 | 2.59E-15 | Up | Same family; supports robust humoral response |
| IGHG4 | +6.02 | 8.07E-14 | Up | Additional IgG subclass upregulation |
| IGHG1 | +5.63 | 1.09E-09 | Up | Broad IgG activation |
| IGKC | +4.71 | 4.03E-12 | Up | Kappa light chain — confirms plasma cell / antibody production |
| FABP6 | -5.82 | 2.81E-10 | Down | Enterocyte / bile acid handling marker — loss suggests epithelial/metabolic dysfunction |
| GSTA1 | -4.26 | 1.72E-09 | Down | Detoxification enzyme — decreased antioxidant/detox capacity |
| RBP2 | -4.81 | 2.32E-08 | Down | Retinol binding — epithelial/mucosal change |
| ENPEP | -3.07 | 1.67E-06 | Down | Aminopeptidase — epithelial/metabolic function |
| MME | -3.22 | 1.85E-06 | Down | Peptidase with roles in peptide processing |
| KCNJ13 | -2.84 | 2.07E-07 | Down | Ion channel — epithelial transport changes |
| LGALS2 | -2.37 | 8.70E-07 | Down | Galectin family — immune/epithelial interactions |

**Figure 2.**
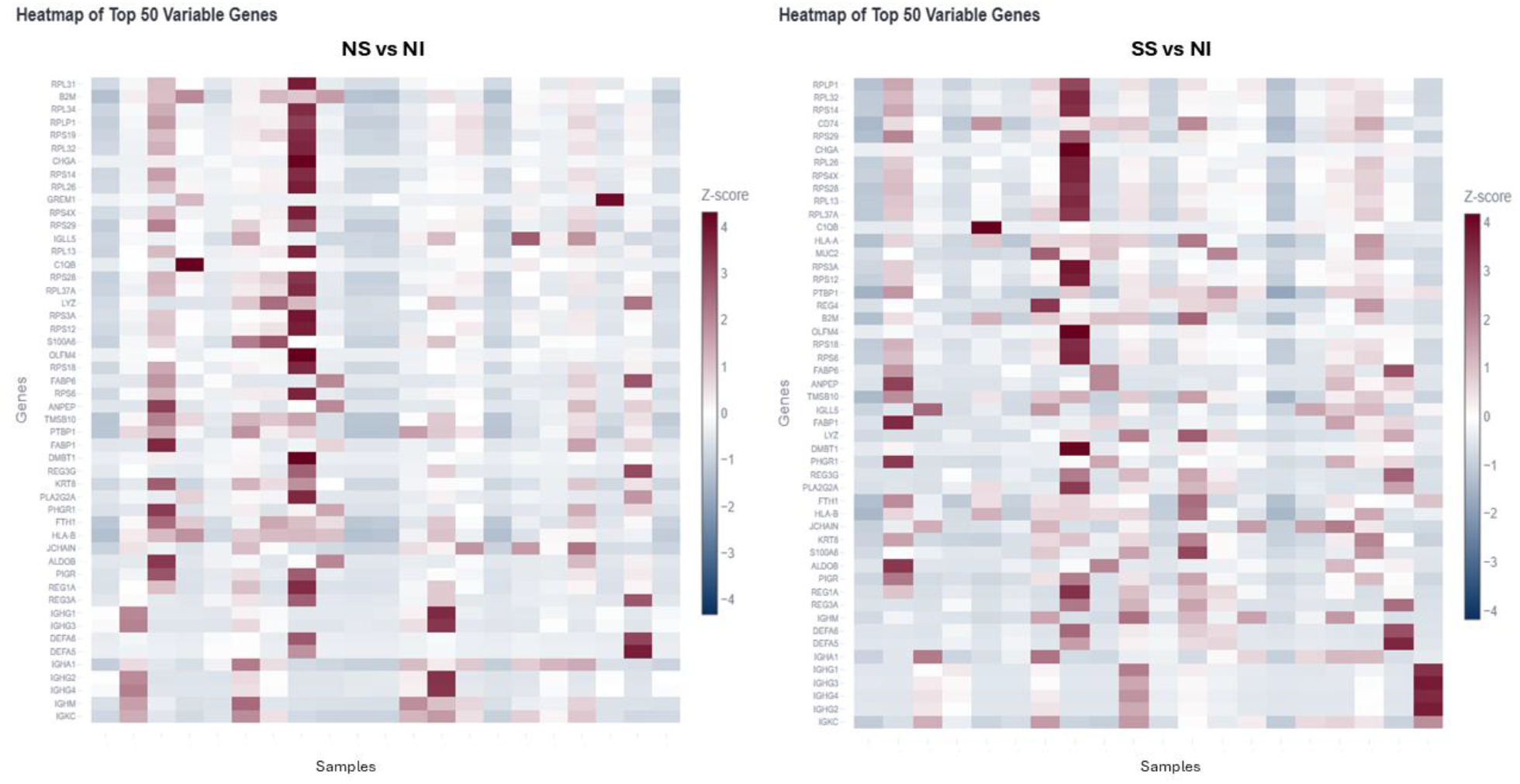
Mucosal DEGs heatmap with top 50 DEGs. Left: NS versus NI; Right: SS versus NI.

### Muscularis Mucosae Region Shows Interfacing Activities and Restricted Stenosis-Associated Divergence

The muscularis mucosae samples demonstrated 122 significant DEGs in NS versus NI and 168 in SS versus NI (Figure 3). Direct SS-versus-NS comparison identified 18 significant DEGs (Table 3), all downregulated, including PLA2G2A, IGHM, SFRP2, REG3A, DEFA5, C7, CCL21, and HLA-DQB1, essentially indicating loss of antimicrobial/epithelial defense gene expression. No significant induction of a broad canonical fibrosis or smooth-muscle proliferation program, except SS versus NI showing decrease of KLF5, GATA6, and MYO7B that potentially play a role in smooth muscle differentiation and function. The SS-versus-NS transcriptional divergence seems to suggest that the muscularis mucosae or peri-muscularis mucosae or muscularis mucosae-submucosa interface may represent a transitional zone in the development of stenosis, although some of the altered genes likely reflect the interfacing basal mucosa and/or superficial submucosa that were inevitably included in the ROIs.

**Table 3.** Significant DEGs in muscularis mucosae in SS-versus-NS transcriptional divergence.

| Gene | log2FC<br>(SS vs NS) | padj | Biologic Significance |
| --- | --- | --- | --- |
| PLA2G2A | -2.16 | 3.80E-05 | Key enzyme in phospholipid metabolism, immune defense, and tissue repair |
| APCDD1 | -1.18 | 2.20E-03 | A membrane-bound Wnt inhibitor, involved in development and tissue homeostasis |
| IGHM | -2.60 | 2.20E-03 | Essential for producing IgM antibodies, the first line of humoral defense |
| C7 | -1.86 | 2.20E-03 | Complement component 7 |
| REG3A | -2.00 | 2.20E-03 | A multifunctional C-type lectin with antimicrobial, regenerative, and signaling roles |
| PDK4 | -1.37 | 2.20E-03 | Pyruvate Dehydrogenase Kinase 4, regulating glucose and fatty acid metabolism |
| SFRP2 | -2.12 | 2.20E-03 | A secreted Wnt pathway modulator with roles in development, tissue homeostasis |
| DEFA5 | -2.18 | 2.29E-03 | Defensin alpha 5, a Paneth cell-specific antimicrobial peptide for mucosal defense |
| CCL21 | -1.66 | 4.02E-03 | A key chemokine that orchestrates immune cell migration and inflammatory responses |
| HLA-DQB1 | -1.75 | 5.53E-03 | Beta chain of DQ heterodimer, enabling antigen presentation to T cells |
| SCN7A | -1.15 | 5.53E-03 | Sodium voltage-gated channel alpha subunit 7 |
| ADH1B | -1.78 | 5.53E-03 | Alcohol dehydrogenase 1B, a key enzyme in ethanol metabolism |
| DEFA6 | -1.89 | 6.23E-03 | A Paneth cell-specific alpha-defensin |
| USP17L11 | -1.16 | 7.66E-03 | A cysteine-type deubiquitinase that participates in protein deubiquitination and regulation of protein stability |
| CFD | -1.51 | 1.44E-02 | Complement Factor D, involved in innate immunity and lipid homeostasis |
| ADAMDEC1 | -1.05 | 4.83E-02 | a secreted metalloproteinase, plays role in dendritic cell function and interactions with germinal center T cells |
| TFPI | -1.03 | 4.83E-02 | Tissue Factor Pathway Inhibitor, a critical natural anticoagulant |
| RNASE1 | -1.20 | 4.83E-02 | Ribonuclease A family member 1, essential for RNA degradation, with roles in immune and coagulation regulation |

**Figure 3.**
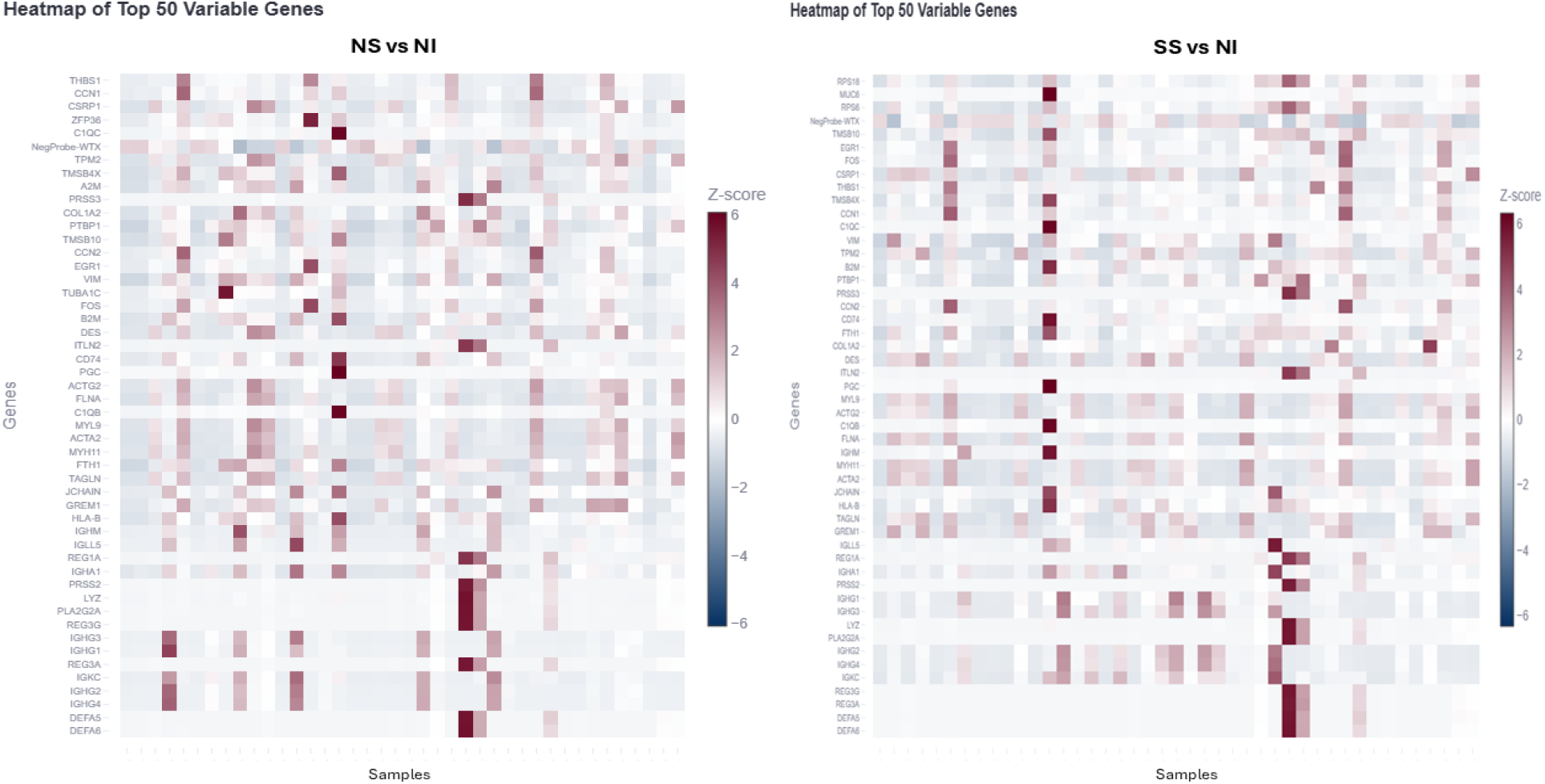
Muscularis mucosae DEGs heatmap with top 50 DEGs. Left: NS versus NI; Right: SS versus NI.

### Mucosa and Submucosa are Transcriptionally Very Different Compartments

In the mucosa-vs-submucosa comparison, all 823 listed genes met padj <0.05, with 156 higher in submucosa and 667 lower. Submucosa is enriched for a stromal/mesenchymal program including SFRP2, C3, C7, CCDC80, GREM1, OGN, FBLN1, MGP, SLIT3, CXCL12, DPT, COL14A1, and MFAP4. In contrast, mucosa is dominated by epithelial and antimicrobial genes such as PIGR, REG3A, REG1A, DEFA5, DEFA6, MUC2, REG3G, LCN2, KRT8, KRT19, and PRSS2. This gave us a very clean molecular definition of the bowel-wall compartments.

### Non-Stenotic Submucosa Shows Inflammatory and Modest Remodeling Programs

Using all histologically valid submucosal ROIs (including some containing lymphoid follicles), NS versus NI identified 132 significant DEGs (131 increased, 1 decreased). The signature was dominated by immunoglobulin/B-cell/plasma-cell biology, including IGHM, IGKC, IGHG1-4, IGHA1, IGLL5, MZB1, XBP1, DERL3, TENT5C, JCHAIN, CD79A, and related genes. In addition, the NS analysis identified a modest stromal/remodeling component, including COL6A3, BGN, COL1A1, COL3A1, THBS1, MMP2, RGS5, and related genes. These findings indicate that the non-stenotic inflamed submucosa is molecularly heterogeneous and is not transcriptionally restricted to humoral inflammation but already has a modest remodeling program. (Figures 4-5, Table 4)

**Table 4.** Selected top submucosal DEGs shared by SS and NS relative to NI.

| Gene | NS vs NI log2FC | NS padj | NS significant | SS vs NI log2FC | SS padj | SS significant | Biologic emphasis |
| --- | --- | --- | --- | --- | --- | --- | --- |
| IGHM | 6.366148362 | 2.46806E-09 | Yes | 4.612097183 | 1.99732E-09 | Yes | Humoral/plasma-cell |
| IGKC | 5.56881652 | 8.05092E-07 | Yes | 5.567516939 | 3.83156E-12 | Yes | Humoral/plasma-cell |
| IGHG1 | 5.059121622 | 0.000392365 | Yes | 5.275091231 | 6.71256E-09 | Yes | Humoral/plasma-cell |
| IGHG2 | 5.525253061 | 4.82059E-06 | Yes | 5.901280051 | 2.11506E-12 | Yes | Humoral/plasma-cell |
| IGHG3 | 5.616531558 | 3.78251E-06 | Yes | 5.905038576 | 1.17254E-12 | Yes | Humoral/plasma-cell |
| IGHG4 | 5.61186146 | 4.82059E-06 | Yes | 6.004711391 | 1.17254E-12 | Yes | Humoral/plasma-cell |
| MZB1 | 3.388922599 | 5.42252E-05 | Yes | 2.760536867 | 0.000105226 | Yes | Humoral/plasma-cell |
| XBP1 | 3.063349203 | 0.0002502 | Yes | 2.85420963 | 2.446E-05 | Yes | Humoral/plasma-cell |
| DERL3 | 2.470843009 | 0.0002502 | Yes | 2.310902381 | 0.000600518 | Yes | Humoral/plasma-cell |
| TENT5C | 2.305703471 | 0.0002502 | Yes | 2.080239682 | 0.001535183 | Yes | Humoral/plasma-cell |
| COL6A3 | 2.035372008 | 0.015258158 | Yes | 1.776076916 | 0.003089274 | Yes | ECM/stromal remodeling |
| BGN | 1.619729281 | 0.024229455 | Yes | 1.592272523 | 0.008643183 | Yes | ECM/stromal remodeling |
| COL1A1 | 1.722333787 | 0.031798978 | Yes | 2.62444811 | 0.000201525 | Yes | ECM/stromal remodeling |
| COL3A1 | 1.821475365 | 0.033054297 | Yes | 2.337463281 | 0.001050982 | Yes | ECM/stromal remodeling |
| THBS1 | 1.83195022 | 0.048479905 | Yes |  |  | No | ECM/stromal remodeling |
| MMP2 | 1.82671438 | 0.049010279 | Yes | 1.805194038 | 0.005154826 | Yes | ECM/stromal remodeling |
| RGS5 | 1.337754188 | 0.034682127 | Yes | 2.133337842 | 0.000215764 | Yes | ECM/stromal remodeling |
| GREM1 | 2.130494234 | 0.081763547 | No | 2.565930702 | 0.00425626 | Yes | ECM/stromal remodeling |
| TNC |  |  | No | 2.250856516 | 0.006347161 | Yes | ECM/stromal remodeling |
| ACTA2 |  |  | No | 3.613742989 | 1.42806E-05 | Yes | Contractile/muscularization |
| TAGLN |  |  | No | 3.273652631 | 0.000452148 | Yes | Contractile/muscularization |
| TPM2 |  |  | No | 2.896399973 | 0.000600518 | Yes | Contractile/muscularization |
| MYL9 |  |  | No | 2.663403635 | 0.001975114 | Yes | Contractile/muscularization |
| MYLK |  |  | No | 2.421631347 | 0.003352001 | Yes | Contractile/muscularization |
| CNN1 |  |  | No | 2.357029367 | 0.006445361 | Yes | Contractile/muscularization |
| MYH11 |  |  | No | 2.326785781 | 0.019580004 | Yes | Contractile/muscularization |
| ACTG2 |  |  | No | 2.124861924 | 0.042394796 | Yes | Contractile/muscularization |
| ITGA8 |  |  | No | 1.966465528 | 0.010405474 | Yes | Cell-matrix adhesion |

**Figure 4.**
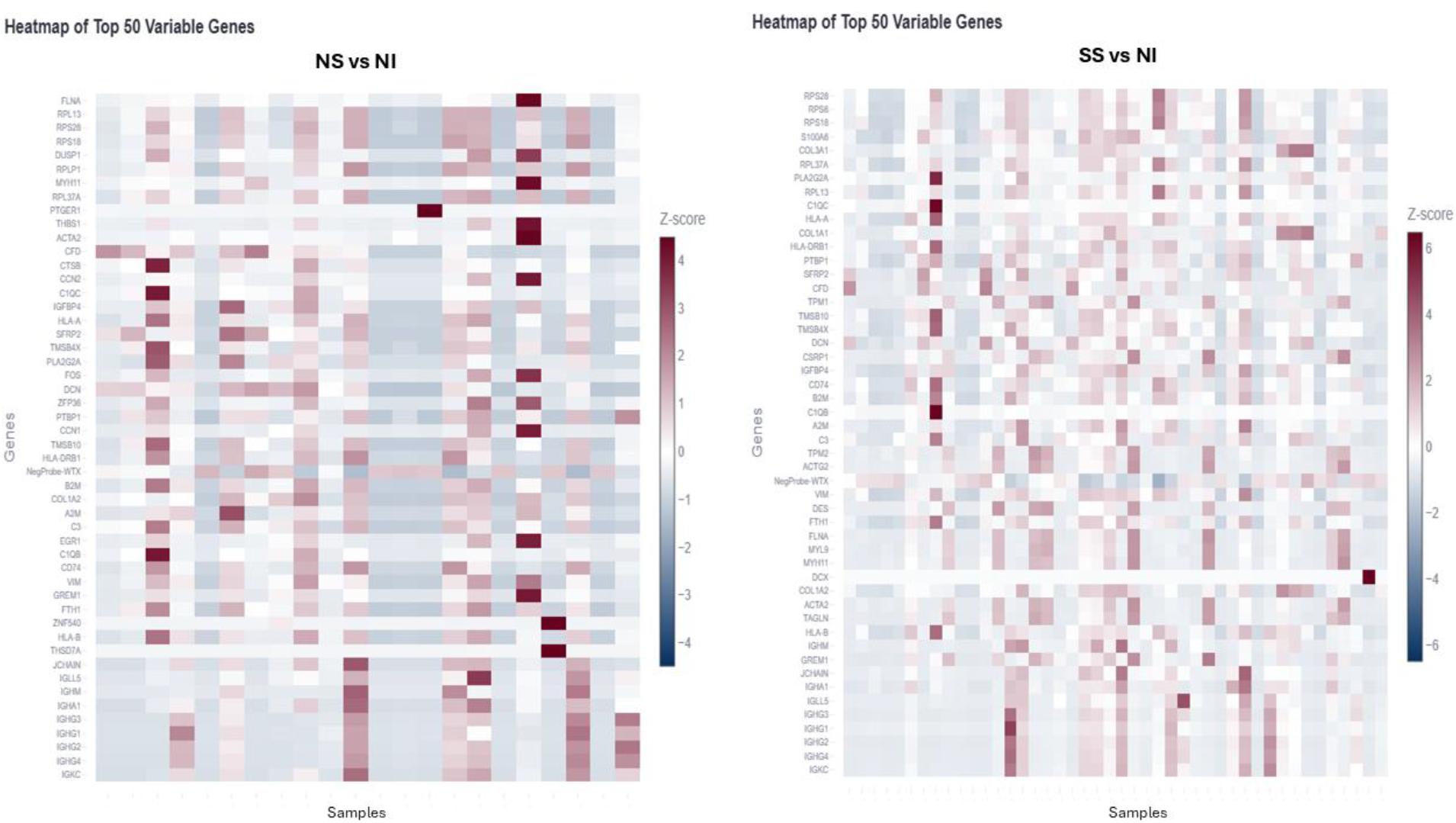
Submucosal DEGs heatmap with top 50 DEGs. Left: NS versus NI; Right: SS versus NI.

**Figure 5.**
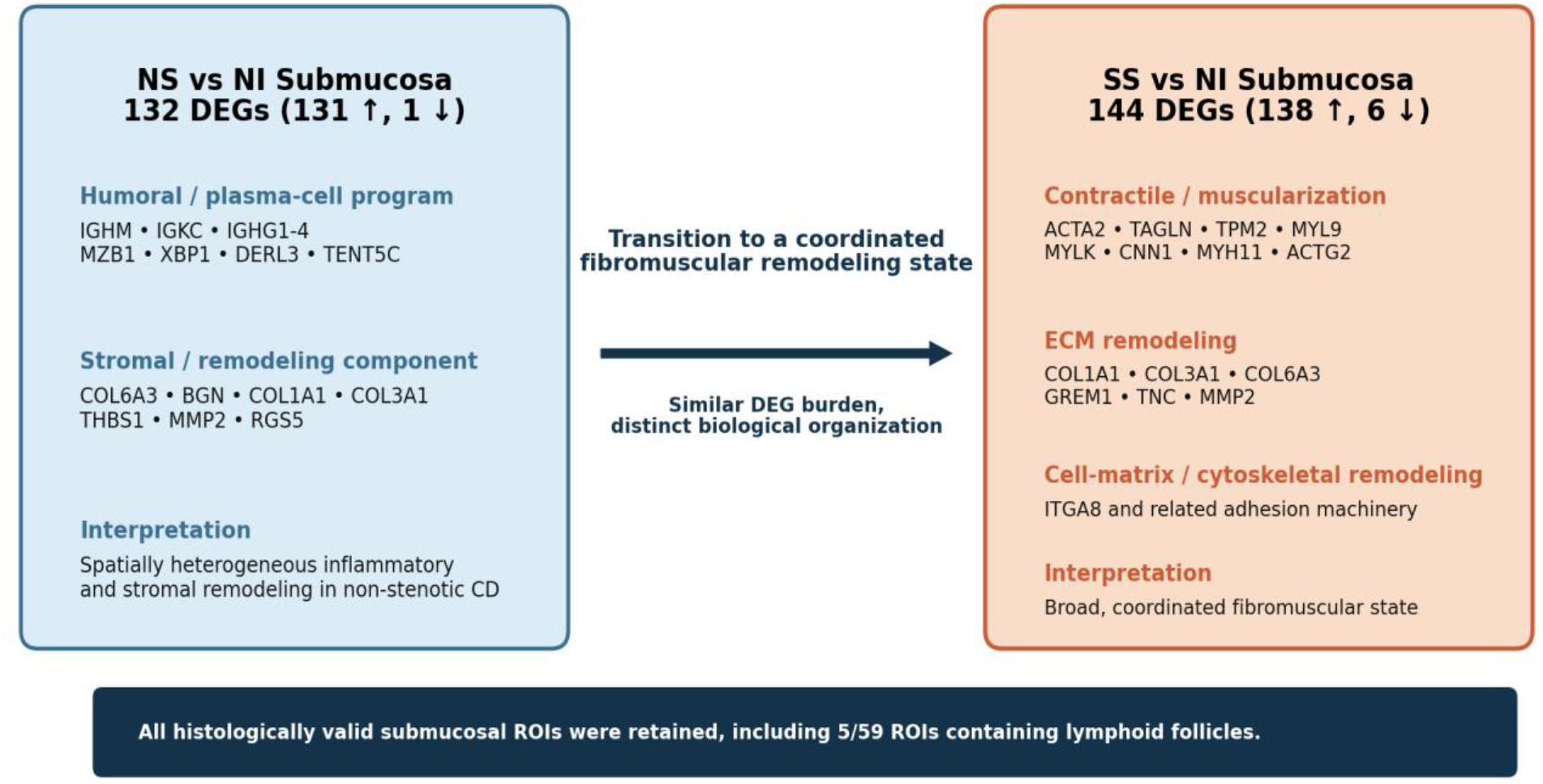
Submucosal transcriptional programs. NS submucosa demonstrates a dominant humoral/plasma-cell response with a modest stromal/remodeling component, whereas SS submucosa demonstrates a broad ECM, contractile, cytoskeletal, and cellmatrix adhesion program. All histologically valid ROIs are included in the primary analysis.

### Stenotic Submucosa Demonstrates a Robust Fibromuscular Remodeling Phenotype

SS versus NI submucosa identified 144 significant DEGs (138 increased, 6 decreased). A prominent contractile/smooth-muscle program included ACTA2, TAGLN, TPM2, MYL9, MYLK, CNN1, MYH11, ACTG2, DES, LMOD1, and CALD1. ECM and remodeling genes included COL1A1, COL3A1, COL6A3, GREM1, TNC, MMP2, LTBP1, BGN, SPARC, and related genes, while cell-matrix/cytoskeletal genes included ITGA8 and additional adhesion-associated components. Thus, the SS submucosa combined inflammatory/humoral activity with a broad and coordinated structural remodeling program. (Figures 4-5, Table 4)

Direct SS-versus-NS comparison identified 43 significant DEGs, the largest direct stenosis-associated difference among the 4 bowel-wall compartments. Together with the absence of significant SS-versus-NS DEGs in mucosa and muscularis propria, this finding localizes the strongest stenosis-associated transcriptional divergence to the submucosa. Of all the 43 DEGs, 42 are downregulated in SS and only one is upregulated: WFDC1 (+1.59, padj=0.0407). A striking proportion of the downregulated genes were canonical histone-family genes including H4C15, H4C13, H3C15, H2AC16, H1-5, H2AC11, H2AC17,

H2BC14, H3C10 and many others, suggesting a reduced proliferative/cell cycle-associated transcriptional component rather than a stenosis-specific signaling pathway. A coordinated downregulation of several other genes is also observed that included NR4A1, CCL18, NAMPT, IRF8, C1QB, NCF1, HLA-DMB, CLDN11, PLA2G2A, and KRT73. The latter findings strongly suggest reduced antigen-presenting and myeloid cell activity, dampened innate/adaptive immune tone, and concurrent remodeling with a shift in spatial composition. NR4A1 is of particular interest, since its loss may suggest a lower fibroblast/SMC activation threshold. NAMPT is known to play role in cell metabolism and survival.

### Fibrotic and Muscularized Submucosa in Fibrostenosis Represent Distinct Transcriptional States

Direct spatial comparison of histologically muscularized versus fibrotic SS submucosa identified 219 significant DEGs, with 159 higher in muscularized regions and 60 higher in fibrotic regions. Muscularized regions showed striking enrichment of DES (+4.72), ACTG2 (+4.42), MYH11 (+4.02), TAGLN (+3.74), CNN1 (+3.61), TPM2 (+3.38), ACTA2 (+3.34), MYL9 (+2.91), MYLK (+2.80), and LMOD1 (+2.22). Remodeling-associated genes enriched in muscularized regions included TNC (+3.17), LTBP1 (+2.35), WFDC1 (+2.35), ITGA8 (+2.08), INHBA (+2.04), and ASPN (+1.67). Genes relatively enriched in fibrotic regions included PLA2G2A, CFD, APOE, FABP4, SFRP1, GPX3, SLIT3, and FBLN2. These findings indicate that histologically fibrotic and muscularized submucosa are molecularly distinguishable remodeling states rather than simply different degrees of one process. (Figure 6)

**Figure 6.**
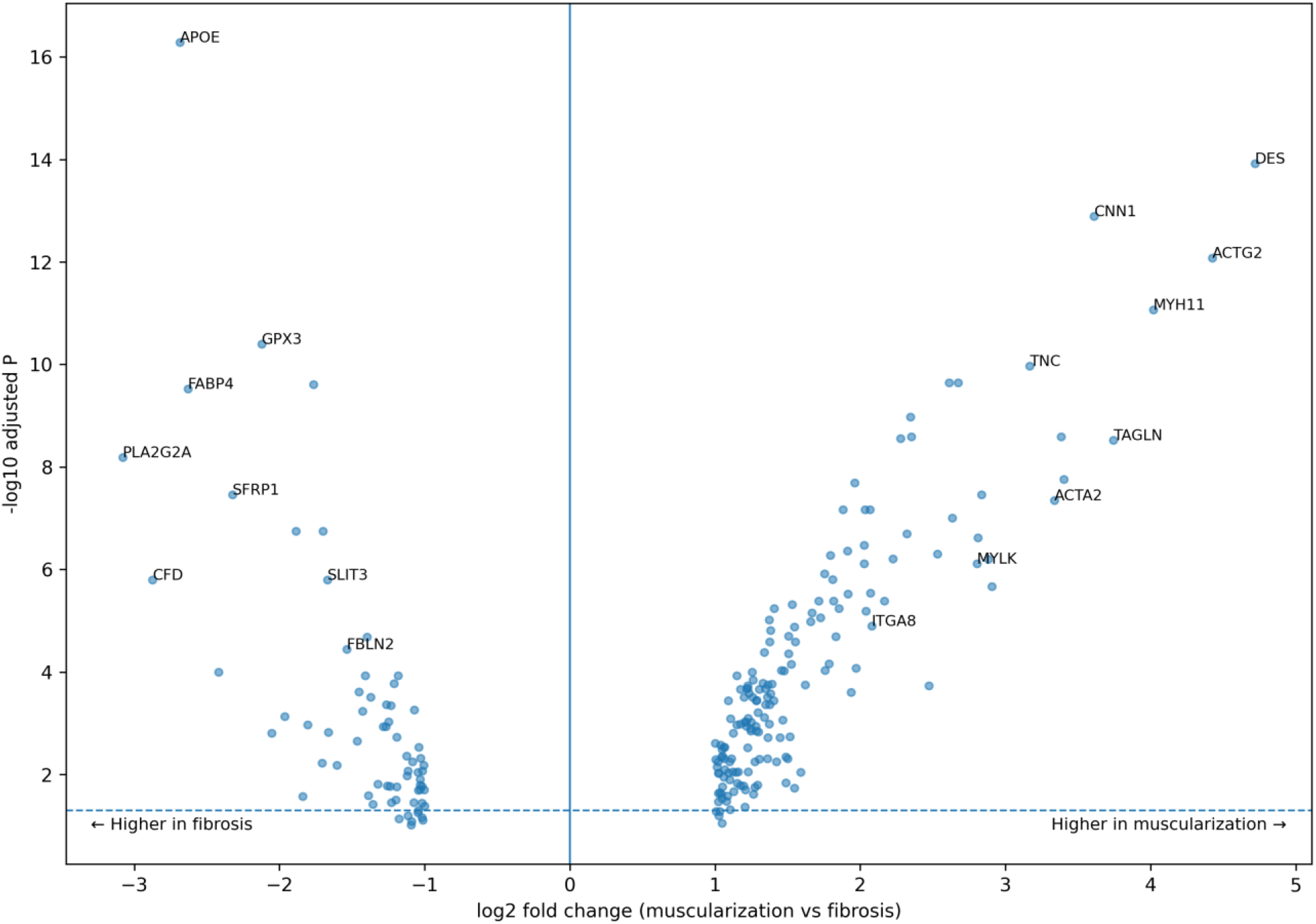
Direct comparison of muscularized versus fibrotic SS submucosa. Positive log2 FC indicates relative enrichment in muscularized regions; negative values indicate relative enrichment in fibrotic regions. Muscularization is characterized by a strong mature smooth-muscle/contractile program.

### Presence of Lymphoid-Follicles in Submucosa Define Distinct Regional Microenvironments

Across submucosal ROIs, lymphoid-follicle-rich (LFR) versus lymphoid-follicle-null (LFN) comparison identified 197 significant DEGs (188 higher and 9 lower in LFR). LFR regions were enriched for an organized lymphoid/B-cell program, including CR2, MS4A1, FCRL1, CD22, FDCSP, CCL19, CXCL13, LTB, CD19, and CD79B. In contrast, the small set of genes significantly lower in LFR included the contractile/fibromuscular genes DES, MYH11, and TAGLN, together with ASPN and MFAP2. Additional contractile and remodeling genes, including ACTA2, ACTG2, CNN1, GREM1, ITGA8, THBS1, TNC, MMP2, and COL1A1, showed coordinated directional shifts toward LFN but did not individually meet the adjusted-P-value criterion.

Comparing LFR areas of submucosa in SS versus that in NS identified 408 significant DEGs (402 higher and 6 lower in SS). SS-LFR regions demonstrated a strong fibroblast/ECM-remodeling program, including GREM1, SFRP2, DCN, COL1A1, COL1A2, COL3A1, COL6A1, COL6A2, COL6A3, MMP2, and TIMP1. Thus, the presence of a lymphoid-follicle-rich microenvironment did not preclude acquisition of a robust stenosis-associated stromal program.

Within SS alone, LFR versus LFN yielded only 1 DEG meeting the significance criteria: CCL19 (log2 fold change, +3.62; padj=4.01E-06), enriched in LFR. Nevertheless, a coordinated but non-FDR-significant directional pattern was evident: lymphoid-associated genes such as FCRL1, MS4A1, CXCL13, and CD22 tended to be higher in LFR, whereas GREM1, SFRP2, multiple collagen/ECM genes, and contractile genes including DES, MYH11, TAGLN, ACTG2, MYLK, CNN1, and ACTA2 tended to be higher in LFN. Because these latter genes did not survive multiple-testing correction in the SS-only comparison, they are interpreted as only an exploratory spatial pattern. (Figure 7)

**Figure 7.**
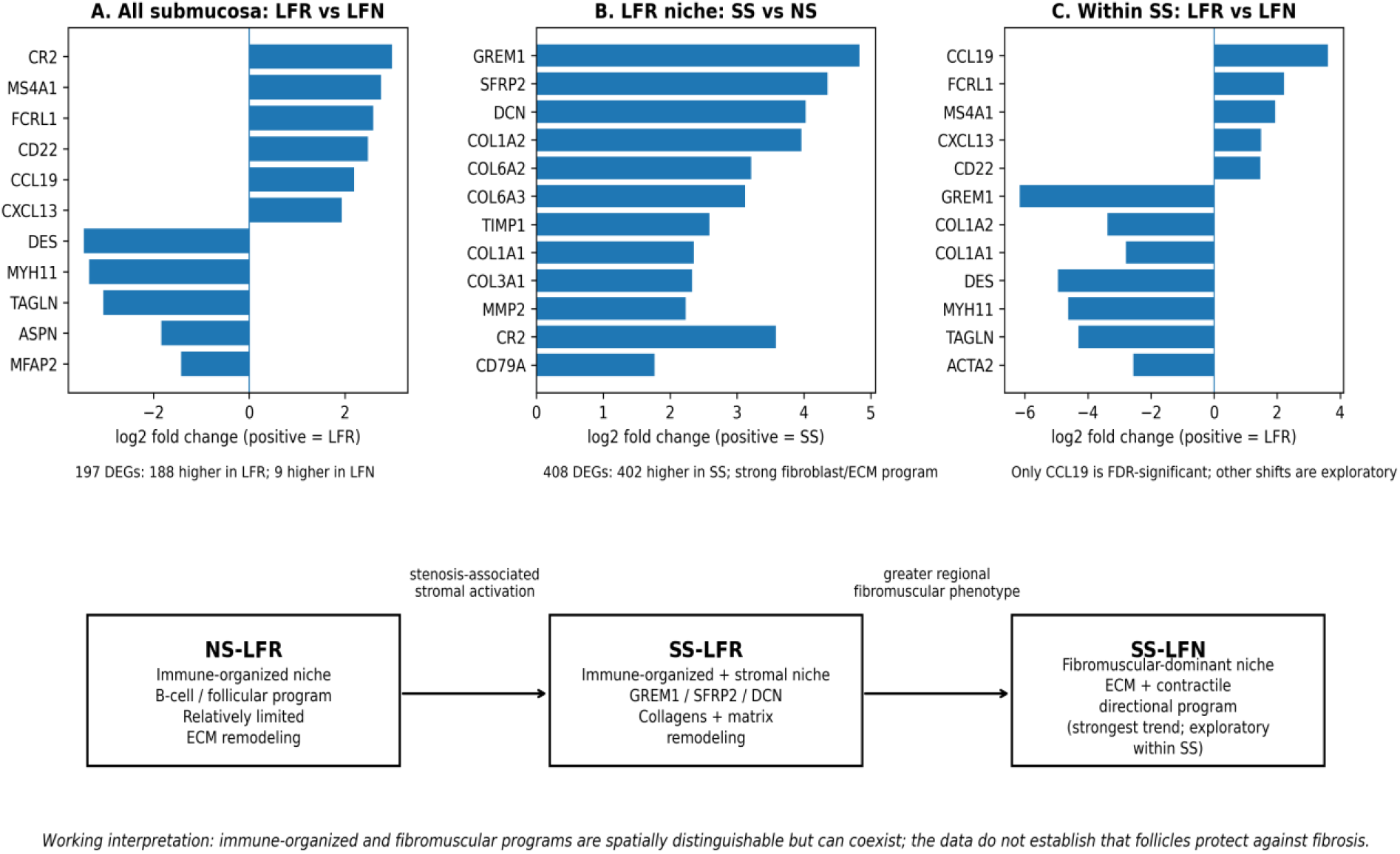
Working model of regional immune-organized and fibromuscular submucosal microenvironments. LFR regions show organized lymphoid/B-cell biology; SS-LFR acquires a fibroblast/ECM-remodeling program; within SS, the strongest fibromuscular directional pattern is observed in LFN regions.

### Sensitivity to Exclusion of Lymphoid-Follicle-Predominant ROIs

Exclusion of the 5 follicle-predominant submucosal ROIs markedly altered significantly the NS-versus-NI result, reducing the number of DEGs from 132 to 23, which is substantially different from the previous analysis that included lymphoid follicles, whereas SS-versus-NI was changed from 144 to 165 significant DEGs. Therefore, in comparison without lymphoid follicles the difference between NS and NS seems to be enormous. However, because the excluded ROIs were patient-clustered (3 NS ROIs from one patient and 2 SS ROIs from another patient), the NS change cannot be attributed specifically to lymphoid-follicle biology and may reflect regional and/or patient-specific influence on the fitted contrast. More importantly, in the corrected submucosa, the NS submucosa shows primarily a humoral/plasma-cell-associated inflammatory program (with dominant genes including IGHG, IGKC, IGHM, IGHA1, IGLL5, MZB1, TXNDC5, XBP1, and CD79A) rather than a strong fibromuscular remodeling program. In contrast, the additional SS-associated genes are heavily enriched for fibromuscular, ECM, contractile, and cell–matrix biology, while the shared humoral/plasma-cell signals were not dismissed. Therefore, the core SS ECM-contractile-cytoskeletal-adhesion programs remained highly stable, supporting the robustness of the established SS fibromuscular phenotype.

### Muscularis Propria Shows Inflammatory-Predominant Changes but No Detectable Classic Fibromuscular Programs

Muscularis propria demonstrated 16 significant DEGs in NS versus NI and 34 in SS versus NI (Figure 8). The SS muscularis propria showed a seemingly stronger disease-associated transcriptional response than NS, but the pattern is dominated by humoral/plasma-cell genes rather than classic smooth-muscle or fibrosis genes. The most prominent SS-vs-NI increases are IGHG4, IGHG2, IGHG3, IGHG1, IGLL5, IGKC, MZB1, IGHM. This confirms a very strong plasma-cell/humoral immune signature extending into the muscularis propria. However, direct SS-versus-NS comparison identified no statistically significant DEGs after multiple-testing correction. IGHM, IGLL5, and TM6SF1 showed absolute log2 fold changes greater than 1 in the displayed comparison, but none remained significant after adjustment (all adjusted P= 1.00). Both NS and SS muscularis propria show marked reductions in DEFA5, DEFA6, REG3A, REG3G, and PLA2G2A. These are not canonical muscularis propria genes, so they may reflect reduced epithelial/Paneth-associated transcript contamination or true changes in adjacent tissue composition rather than intrinsic muscle biology.

**Figure 8.**
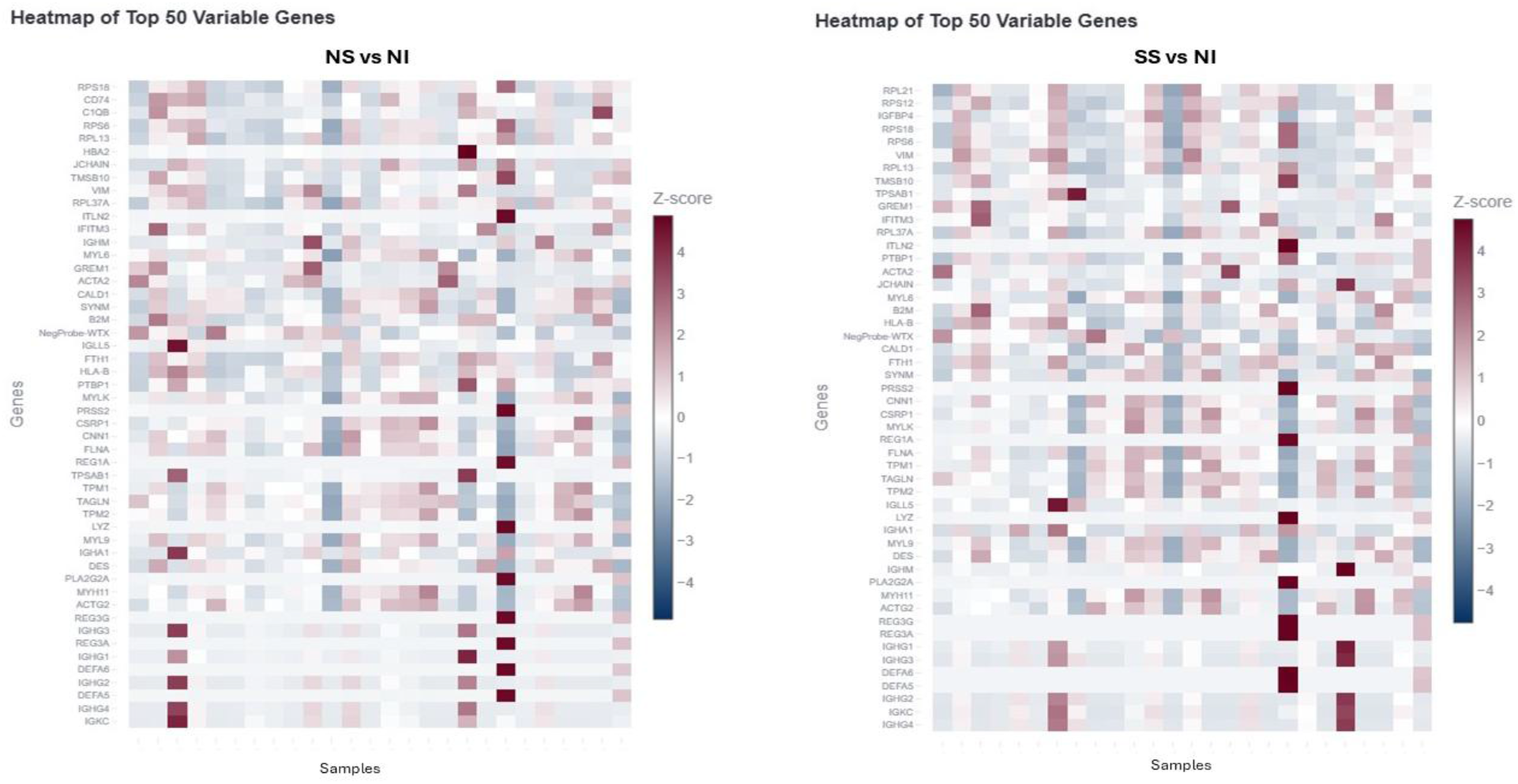
Muscularis propria DEGs. Left: NS versus NI. Right: SS versus NI.

At the same time, several interesting non-immunoglobulin genes were significantly increased in SS versus NI comparison: PIM2, SIK1, TPSAB1, POSTN, TXNDC5, PKN3, and COL14A1 (log2FC +1.33 to +2.41). The increase of PIM2 and SIK1 is potentially signaling smooth muscle cell growth and survival and hence contributes to smooth muscle hypertrophy in muscularis propria; however, it is uncertain since the SS-versus-NS contrast was not significant. Additionally, POSTN, a classic matrix/remodeling-associated gene, was increased in both NS and SS muscularis propria, with a somewhat larger effect in SS. TXNDC5 was also increased in both groups but more strongly in SS. The pattern appears to be compatible with a progressive stromal/remodeling response; however, we did not see a significant difference in SS-versus-NS analysis to confirm. Moreover, upregulated NOTCH3 and XBP1 in similar NS-versus-NI and SS-versus-NI comparison are noteworthy for their possible roles in the maintenance and differentiation of neuronal and glial cells in the enteric nervous system in the muscular layer. Overall, unlike the submucosa, muscularis propria did not demonstrate a significantly detectable classic fibromuscular transcriptional program and appeared less transcriptionally distinctive for fibrostenosis than the submucosa.

## Discussion

This histomorphology/molecular marker-ROI guided layer-by-layer and disease stage-resolved spatial transcriptomic study identifies a compartmentalized molecular architecture of fibrostenotic CD. The principal finding is that stenosis is associated with acquisition of a broad submucosal fibromuscular remodeling program rather than simply amplification of mucosal inflammation. The all-inclusive submucosal dataset, which retained all histologically valid ROIs including the small number containing prominent lymphoid follicles, also reveals an important nuance: non-stenotic inflamed submucosa is molecularly heterogeneous and can already display modest stromal/remodeling features.

Inflammation creates the permissive disease environment, but it does not by itself explain why only one short segment becomes stenotic. A major challenge in CD fibrosis research is separating molecular abnormalities associated with chronic inflammation from those specifically associated with structural stenosis. Both NS and SS mucosa demonstrate marked inflammatory, immunoglobulin, and epithelial/metabolic abnormalities relative to NI, yet direct SS-versus-NS analysis identifies no significant mucosal DEGs. Thus, prominent mucosal inflammation is shared across diseased states and does not by itself account for stenosis.

The submucosa emerges as the dominant remodeling compartment harboring phenotype-defining stenotic alterations. Direct SS-versus-NS comparisons identify 0 significant DEGs in mucosa, 18 in muscularis mucosae, 43 in submucosa, and 0 in muscularis propria. In the all-inclusive submucosal analysis, NS versus NI contains a strong humoral/plasma-cell program together with a more limited stromal signature that includes COL1A1, COL3A1, COL6A3, BGN, THBS1, MMP2, and RGS5. SS versus NI, however, demonstrates a much broader coordinated program involving ECM deposition/remodeling, cell-matrix adhesion, cytoskeletal organization, and mature contractile machinery. This pattern supports a continuum in which stromal remodeling may be regionally present before fixed stenosis, while established SS is characterized by a more extensive fibromuscular tissue state.

The muscularis mucosae are usually treated as an anatomical boundary between mucosa and submucosa. In our data the muscularis mucosa layer showed further suppression of epithelial/innate-defense, immune, metabolic genes as well as stromal-associated transcriptional divergence but no induction of a broad canonical fibrosis or smooth-muscle proliferation program. The findings partly reflect the changes in the interfacing mucosa and submucosa and suggest that the muscularis mucosae-submucosa interface may represents a transitional zone with molecular transition/dysregulation preceding or adjacent to the much stronger fibromuscular phenotype in the deeper tissue. In view of the common histopathological finding of downward dissection of smooth muscle cells within muscularis mucosae in chronically diseased bowel, concurrent smooth muscle cell migration from muscularis mucosa into submucosa is also possible.

Our layer-resolved findings are concordant with recent single-cell work showing that many stricture-selective transcriptional changes occur in the mucosa/submucosa rather than the muscle layer.^8^ They also complement recent single-cell/spatial studies identifying IgG-positive plasma cells, inflammatory fibroblasts, collagen-high fibroblasts, and spatially organized stromal-immune networks in stricturing CD.^9,13,14^ The present study adds a pathology-defined comparison across bowel-wall layers and directly resolves histologically fibrotic from muscularized stenotic submucosal regions.

Crohn’s stricture is a fibromuscular lesion. The traditional term ‘fibrostenotic’ appropriately recognizes fibrosis but may underemphasize the contribution of smooth-muscle remodeling. ACTA2, TAGLN, MYH11, ACTG2, CNN1, MYLK, DES, TPM2, and MYL9 are increased in SS submucosa, and direct comparison of muscularized with fibrotic SS regions produces even larger effect sizes for many of these genes. These findings provide molecular support for the histopathologic observation that muscularization contributes substantially to bowel-wall thickening. Luminal narrowing may therefore reflect at least two interrelated structural processes: ECM accumulation and contractile-cell expansion/remodeling. This distinction may have therapeutic implications because targeting collagen deposition alone may not reverse an established muscularized component of a stricture.

Fibrosis and muscularization are related but molecularly distinct. Muscularized SS regions are characterized by exceptionally high expression of highly differentiated smooth muscle contractile gene programs together with TNC, LTBP1, ITGA8, INHBA, and other remodeling-associated genes, whereas fibrotic regions show relative enrichment of APOE, FABP4, GPX3, SFRP1, CFD, SLIT3, and FBLN2. Together these findings support a model in which fibrosis and muscularization represent related but transcriptionally distinct remodeling states, rather than a single process in which progessive ECM deposition culminates in muscularization. A better hypothesis is that the stenotic submucosa contains coexisting remodeling niches representing different stromal/mesenchymal states or remodeling trajectories. The present cross-sectional data cannot establish their lineage relationship or temporal sequence in the mixed process; that question should be addressed by future single-cell, higher-resolution spatial, or lineage-oriented studies.

The concurrent increase of ITGA8, ITGB1, TLN1, FERMT2, PARVA, VCL, and TNS1, together with focal-adhesion pathway enrichment, suggests that altered cell-matrix attachment accompanies the contractile phenotype. These findings provide a plausible mechanistic framework in which ECM remodeling and contractile-cell behavior are mechanistically linked (Figure 9). However, because the present study measures transcript abundance rather than pathway activation or tissue mechanics, this should be considered a putative cell-matrix-adhesion-contractility axis rather than demonstrated ECM-stiffness signaling alone. Future validation could examine integrin-FAK-SRC-RHOA/cytoskeletal remodeling pathway activity in relation to α-SMA/MYH11-positive muscularized regions.

**Figure 9.**
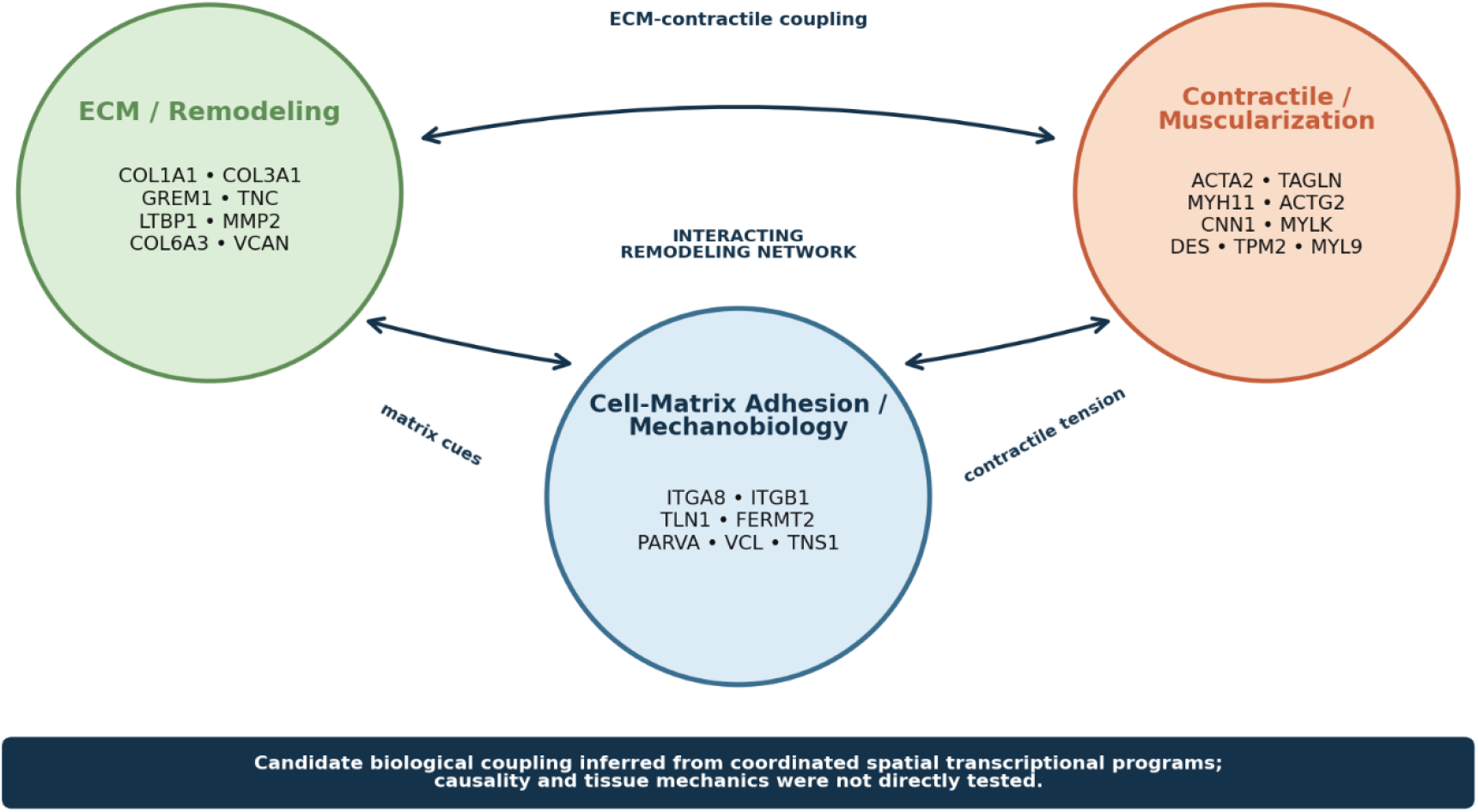
Integrated model of submucosal fibromuscular remodeling in Crohn’s stenosis. The transcriptomic data demonstrate coordinated ECM/remodeling, cell-matrix adhesion, and contractile gene programs. Bidirectional arrows indicate proposed biological interaction and do not establish causal direction.

GREM1 and TNC remain particularly attractive candidate markers of active fibromuscular remodeling. GREM1 is significantly increased in SS submucosa, while TNC is increased in SS and strongly enriched in muscularized relative to fibrotic SS regions. Independent transcriptomic studies have identified GREM1 as a stricture-associated transcript and localized its expression primarily to fibroblasts.^13^ These findings support prioritizing GREM1, TNC, and ACTA2/MYH11 for future spatial protein validation. In combination with some related structural genes, these candidates may provide a more informative signature of active muscularizing/fibromuscular remodeling than conventional collagen markers alone.

SFRP2 should be interpreted as a stromal-state marker rather than simply a stenosis marker. Its behavior varies according to anatomic and histologic context; it is reduced in the muscularis mucosae SS-versus-NS comparison and does not behave like a simple universal stenosis-upregulated marker. SFRP2 may therefore be more useful for distinguishing stromal states or fibroblast compartments than as a stand-alone fibrostenosis biomarker.

Presence of prominent submucosal lymphoid follicles (LFs) is common in Crohn’s disease, including within stenotic regions. Whether the LFs may contribute to the development of fibrostenosis in CD is unknown. Our candid analyses about LFR/LFN indicate that lymphoid-follicle content contributes to regional transcriptional heterogeneity within the submucosa, although its effects cannot be fully separated from the patient-specific variation in ROI composition. Across submucosal ROIs, LFR regions were characterized by a strong organized lymphoid/B-cell program, whereas several contractile/fibromuscular genes were relatively enriched in LFN regions. Despite their prominent lymphoid transcriptional profile, LFR regions retained evidence of stromal remodeling, particularly in stenotic regions (or tissue). When LFR regions were compared directly between disease states, SS-LFR showed a striking fibroblast/ECM program relative to NS-LFR, including GREM1, SFRP2, DCN, fibrillar and type VI collagens, and matrix-remodeling genes. Thus, a lymphoid-organized niche can coexist with substantial stromal remodeling once fibrostenotic disease is established. These observations underscore the spatial heterogeneity of NS while demonstrating that the SS remodeling signature remains stable despite variation in the lymphoid follicle signature defined by the ROI composition. Within SS, CCL19 was the only LFR-versus-LFN gene to remain significant after multiple-testing correction, strongly supporting organized lymphoid identity of the LFR niche. At the same time, coordinated non-FDR-significant shifts of GREM1/SFRP2, collagen/ECM genes, and mature contractile genes toward LFN suggest a possible spatial gradient in which the most pronounced fibromuscular phenotype preferentially localizes outside follicle-rich regions. This pattern should not be interpreted as evidence that lymphoid follicles promote or suppress stromal remodeling. A more conservative model is that Crohn’s submucosa contains overlapping but distinguishable immune-organized and fibromuscular microenvironments, and that fibrostenosis is accompanied by stromal remodeling even within the immune-organized niche.

The muscularis propria findings further sharpen the compartment-specific interpretation. Although disease-associated changes are present in NS-versus-NI and SS-versus-NI comparisons, direct SS-versus-NS analysis yields no significant DEGs after multiple-testing correction. The absence of a detectable stenosis-specific signature in the muscularis propria, together with the strong submucosal remodeling program supports a compartmentalized model in which the principle transcriptional alteration associated with established stenosis are concentrated (or localized) within the submucosa rather than uniformly distributed across the bowel wall. In view of the common finding of muscularis propria thickening, which has been considered as hypertrophy, it is somewhat puzzling that no significant upregulation of genes directly related smooth muscle cell biology was revealed.

The compartmental distribution of these transcriptomic changes may have clinical implications. Routine endoscopic biopsies primarily sample mucosa, whereas the strongest stenosis-associated remodeling program in this study resides within the submucosa. This raises the possibility that mucosal biomarkers may incompletely capture the structural and molecular biology responsible for established stenosis. Future biomarkers of fibrostenosis may therefore need to reflect deeper-wall remodeling through circulating matrix products, imaging correlates, advanced endoscopic sampling, or molecular signatures linked specifically to fibromuscular remodeling. This remains a hypothesis rather than a clinical conclusion from the current discovery cohort.

These findings should be interpreted in the context of several study-design limitations. First, the cohort comprised only four patients with established fibrostenotic phase of disease requiring surgical resection, which limits generalizability to earlier stages of stricture development. Second, multiple ROIs per patient increased spatial sampling but did not constitute independent biological replication, although patient identity was included as a blocking factor in the differential-expression models. Third, GeoMx ROI-level profiling provided regional spatial resolution rather than single-cell resolution, and histologically defined ROIs may contain heterogeneous cell populations. Regional histologic heterogeneity, including lymphoid follicles, may influence individual contrasts, as illustrated by the NS sensitivity analysis. Finally, the cross-sectional design precludes inference of temporal sequence or causality, and transcript abundance alone does not establish protein expression, pathway activation, or tissue mechanics.

In conclusion, as summarized and envisaged in Figure 10, Crohn’s fibrostenosis is characterized by a spatially concentrated submucosal fibromuscular remodeling program that distinguishes stenotic from non-stenotic disease. The all-inclusive submucosal data further suggest that NS is heterogeneous and may contain limited regional stromal remodeling before fixed stenosis, whereas established SS shows a broader and more robust ECM-contractile-adhesion phenotype. Transition from non-stenotic inflammation to stenotic stricture is associated with acquisition of a broader submucosal fibromuscular remodeling program rather than simply persistence or amplification of mucosal inflammation. Spatial separation of fibrotic and muscularized molecular states supports a model of Crohn’s stricture as a fibromuscular lesion composed of related but distinct remodeling niches and nominates GREM1/TNC-associated remodeling, cell-matrix adhesion, and contractile pathways for future validation as candidate biomarkers and therapeutic targets.

**Figure 10.**
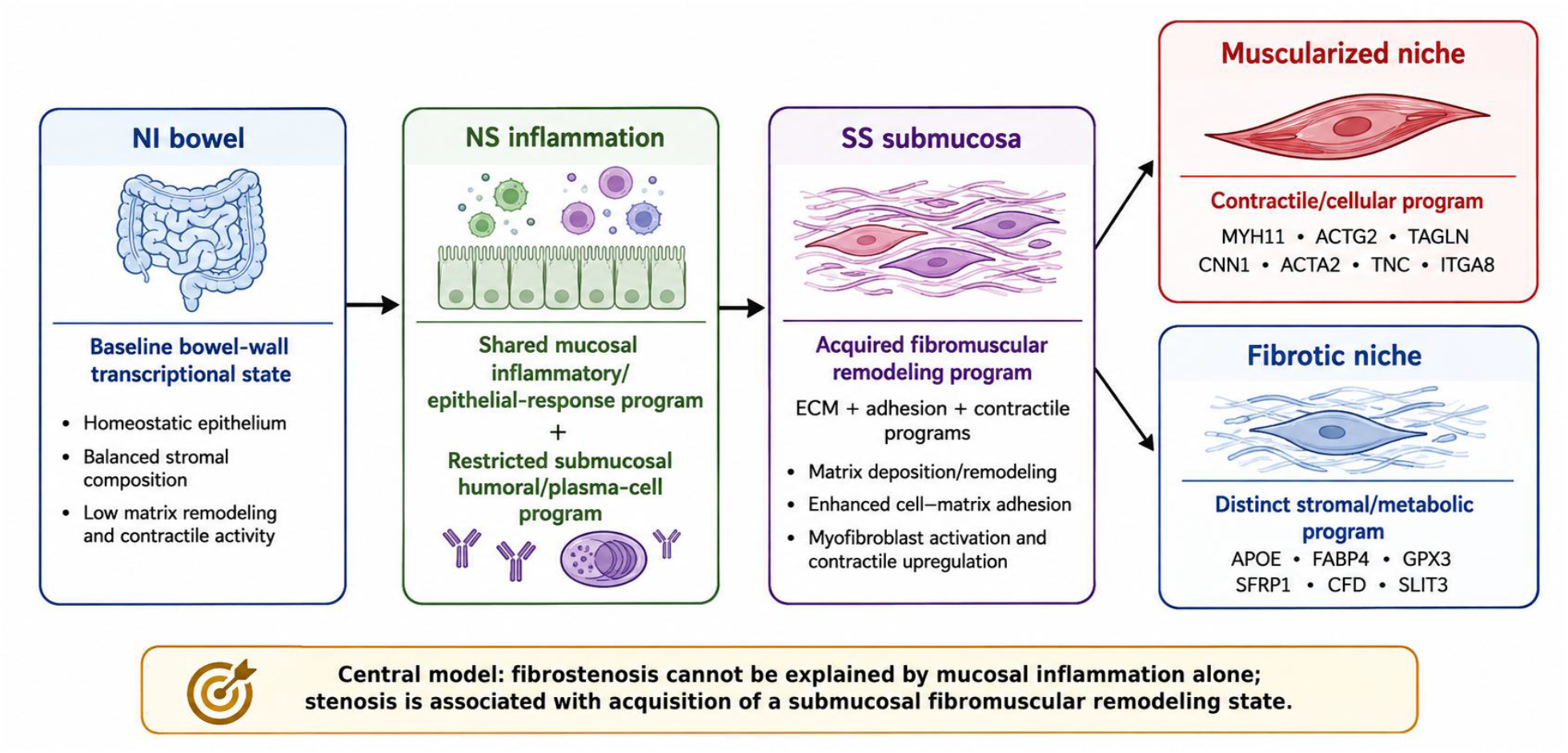
Working model of CD-associated fibrostenotic remodeling. Shared mucosal inflammatory abnormalities characterize NS and SS, while NS submucosa shows heterogeneous humoral and limited stromal remodeling and established SS acquires a broad fibromuscular program with transcriptionally distinguishable fibrotic and muscularized niches.

## Data Availability

All data produced in the present study are available upon reasonable request to the authors

## Abbreviations

CD: Crohn’s disease
DEG: differentially expressed gene
DSP: Digital Spatial Profiler
ECM: extracellular matrix
FFPE: formalin-fixed, paraffin-embedded
NI: non-inflamed
NS: non-stenotic inflamed
ROI: region of interest
SS: stenotic stricture
WTA: Whole Transcriptome Atlas.

## Acknowledgments

Lisa O’Donnell in the Comparative Pathology Laboratory (CPL) performed all the laboratory bench work. CPL is supported in part by NCI Cancer Center Support Grant (P30 CA012197) and North Carolina Biotechnology Center Grant (2015-IDG-1006). Dr. Shuo Niu participated in initial study planning.

## Author Contributions

**XG** concept and proposal development, case materials collection, histopathological evaluation, data analysis, results interpretation, and manuscript writing

**DC** proposal development, DSP laboratory work, data handling, results analysis, and manuscript writing

**GJ** proposal development, DEG data analysis, bioinformatics analysis, AI-guided DESeq2 enabled by CONVERGE-AI, and manuscript writing

## Data Availability Statement

All data is available in the manuscript. Raw data, detailed protocols, and CONVERGE-AI platform in the main text are available upon request to the authors

## References

1. Li C, Kuemmerle JF. Mechanisms that mediate the development of fibrosis in patients with Crohn’s disease. Inflamm Bowel Dis 2014;20:1250–1258. PMID: 24831560.

2. Andoh A, Nishida A. Molecular basis of intestinal fibrosis in inflammatory bowel disease. Inflamm Intest Dis. 2022;7:119–127. PMID: 37064539.

3. Fousekis FS, Mpakogiannis K, Mastorogianni IN, et al. Intestinal fibrosis in Crohn’s disease: pathophysiology, diagnosis, and new therapeutic targets. J Clin Med 2025;14:4060. PMID: 40565806.

4. Koukoulis G, Ke Y, Henley JD, et al. Obliterative muscularization of the small bowel submucosa in Crohn disease: a possible mechanism of small bowel obstruction. Arch Pathol Lab Med 2001;125:1331–1334. PMID: 11570909.

5. Zhang X, Ko HM, Torres J, et al. Luminally polarized mural and vascular remodeling in ileal strictures of Crohn’s disease. Human Pathol 2018;79:42–49. PMID: 29555578.

6. Chen W, Lu C, Hirota C, et al,> et al. Smooth muscle hyperplasia/hypertrophy is the most prominent histological change in Crohn’s fibrostenosing bowel strictures: a semiquantitative analysis by using a novel histological grading scheme. J Crohns Colitis 2017;11:92–104. PMID: 27364949.

7. Gordon IO, Bettenworth D, Bokemeyer A, et al. International consensus to standardise histopathological scoring for small bowel strictures in Crohn’s disease. Gut 2022;71:478–486. PMID: 33952604.

8. Mukherjee PK, Nguyen QT, Li J, et al. Stricturing Crohn’s disease single-cell RNA sequencing reveals fibroblast heterogeneity and intercellular interactions. Gastroenterol 2023;165:1180–1196. PMID: 37507073.

9. Kong L, Subramanian S, Segerstolpe A, et al. Single-cell and spatial transcriptomics of stricturing Crohn’s disease highlights a fibrosis-associated network. Nat Genet 2025;57:1742–1753. PMID: 40562913.

10. Humphreys DT, Lewis A, Pan-Castillo B, et al. Single cell sequencing data identify distinct B cell and fibroblast populations in stricturing Crohn’s disease. J Cell Mol Med 2024;28:e18344.

11. Massimino L, Parigi TL, Riva M, et al. Spatiotemporal analysis of Crohn’s disease reveals PECAM2 signaling at the basis of the inflammation-to-fibrosis transition. J Crohn’s Colitis 2025;19:jjaf130.

12. Sun S, Wang J, Li K, et al. Elucidating the role of IgA plasma cells and PECAM1-CD38 interaction in intestinal fibrosis: a single-cell RNA sequencing analysis in Crohn’s disease. BMJ Gastroenterol 2025;25:712.

13. Acharjee A, Shivaji U, Santacroce G, et al. Novel transcriptomic signatures in fibrostenotic Crohn’s disease: dysregulated pathways, promising biomarkers, and putative therapeutic targets. Inflamm Bowel Dis 2025;31:1502–1513. PMID: 39977234.

14. Zhang D, Zou X, He M, et al. Elucidating a myofibroblast-dominated fibrotic niche in Crohn’s disease-associated fibrostenosis through high-resolution spatial transcriptomics. Cell Mol Gastroenterol Hepatol 2026;20:101701. PMID: 41344440.

15. Kalafateli M, Tourkochristou E, Tsounis EP, et al. New insights into the pathogenesis of intestinal fibrosis in inflammatory bowel diseases: focusing on intestinal smooth muscle cells. Inflamm Bowel Dis 2025;31:579–592. PMID: 39680685.

16. Alfredsson J, Wallem CS, Ostling M, et al. Tissue-layer-resolved proteome landscape of Crohn’s disease strictures highlights potential drivers of fibrosis progression. JCI Insight 2026 Feb 10;11(6):e202461. doi:10.1172/jci.insight.202461. PMID: 41665933.

17. Ke BJ, Abdurahiman S, Biscu F, et al. Intercellular interaction between FAP+ fibroblasts and CD150+ inflammatory monocytes mediates fibrostenosis in Crohn’s disease. J Clin Invest 2024;134:e173835. PMID: 39042469.

18. Zhang Y, Wang J, Sun H, et al. TWIST1+FAP+ fibroblasts in the pathogenesis of intestinal fibrosis in Crohn’s disease. J Clin Invest 2024;134:e179472.

